# Diagnostic accuracy of chest X-ray interpretation for tuberculosis by three artificial intelligence-based software in a screening use-case: an individual patient meta-analysis of global data

**DOI:** 10.1101/2022.01.24.22269730

**Authors:** Sandra V. Kik, Sifrash M. Gelaw, Morten Ruhwald, Rinn Song, Faiz Ahmad Khan, Rob van Hest, Violet Chihota, Nguyen Viet Nhung, Aliasgar Esmail, Anna Marie Celina Garfin, Guy B. Marks, Olga Gorbacheva, Onno W. Akkerman, Kgaugelo Moropane, Le Thi Ngoc Anh, Keertan Dheda, Greg J. Fox, Nina Marano, Knut Lönnroth, Frank Cobelens, Andrea Benedetti, Puneet Dewan, Stefano Ongarello, Claudia M. Denkinger

## Abstract

**Background:** Chest X-ray (CXR) screening is a useful diagnostic tool to test individuals at high risk of tuberculosis (TB), yet image interpretation requires trained human readers who are in short supply in many high TB burden countries. Therefore, CXR interpretation by computer-aided detection software (CAD) may overcome some of these challenges, but evidence on its accuracy is still limited.

We established a CXR library with images and metadata from individuals and risk groups that underwent TB screening in a variety of countries to assess the diagnostic accuracy of three commercial CAD solutions through an individual participant meta-analysis.

**Methods and findings:** We collected digital CXRs and demographic and clinical data from 6 source studies involving a total of 2756 participants, 1753 (64%) of whom also had microbiological test information. All CXR images were analyzed with CAD4TB v6 (Delft Imaging), Lunit Insight CXR TB algorithm v4.9.0 (Lunit Inc.), and qXR v2 (Qure.ai) and re-read by an expert radiologist who was blinded to the initial CXR reading, the CAD scores, and participant information. While the performance of CAD varied across source studies, the pooled, meta-analyzed summary receiver operating characteristic (ROC) curves of the three products against a microbiological reference standard were similar, with area under the curves (AUCs) of 76.4 (95% CI 72.1-80.3) for CAD4TB, 83.3 (95% CI 78.4-87.2) for Lunit, and 76.4 (95% CI 72.1-80.3) for qXR. None of the CAD products, or the radiologists, met the targets for a triage test of 90% sensitivity and 70% specificity. At the same sensitivity of the expert radiologist (94.0%), all CAD had slightly lower point estimates for specificity (22.4% (95% CI 16.9-29.0) for CAD4TB, 34.6% (95% CI 25.3-45.1) for qXR, and 41.0% (95% CI 30.1-53.0) for Lunit compared to 45.6% for the expert radiologist). At the same specificity of 45.6%, all CAD products had lower point estimates for sensitivity but overlapping CIs with the sensitivity estimate of the radiologist.

**Conclusions:** We showed that, overall, three commercially available CAD products had a reasonable diagnostic accuracy for microbiologically confirmed pulmonary TB and may achieve a sensitivity and specificity that approximates those of experienced radiologists. While threshold setting and cost-effectiveness modelling are needed to inform the optimal implementation of CAD products as part of screening programs, the availability of CAD will assist in scaling up active case finding for TB and hence contribute to TB elimination in these settings.

## Introduction

Finding efficient and novel ways to diagnose and treat all people with active pulmonary tuberculosis (TB) is crucial to achieving the ambitious United Nations Sustainable Development Goals to end the TB epidemic by 2030. It is estimated that every year worldwide 10 million people fall ill with TB, approximately 2.9 million of whom are not notified or diagnosed (1). Reasons for this large proportion of undiagnosed TB cases are manifold. Individuals with TB may be asymptomatic and therefore not seek care, but they may also receive irrelevant or inaccurate tests for TB, leaving them undiagnosed. Targeted screening of individuals who are at increased risk of having TB is proposed by the World Health Organization (WHO) as a strategy to detect TB cases earlier (2).

Easy, low-cost, and highly sensitive screening tests are needed for such strategies to quickly rule out TB in the vast majority of screened individuals, so that further highly accurate confirmatory testing can then carried out on those who screened positive (3).

Chest X-ray (CXR) interpreted by a trained human reader has been recognized by WHO as an initial screening test given its high sensitivity (2,4), it can also identify TB in patients who do not report classical TB symptoms (5). However, lack of access to well-trained human readers, and the fact that X-ray equipment is most often available only at district-level centers and referral hospitals, hinders wide-scale use of CXR screening at the community level in many high TB burden countries (6).

In recent years, there has been a growth in the development of computer-aided detection (CAD) software – artificial intelligence-based deep learning algorithms that can interpret digital CXRs. Several CAD products, trained on millions of images, are now capable of detecting TB by producing an abnormality score for TB that can triage individuals who require further confirmatory testing (7,8). Furthermore, CAD can create a heatmap or annotated image indicating on each X-ray image the location of the abnormal findings. Some commercial CAD products, such as CAD4TB (Delft Imaging, Netherlands), are designed for TB detection only, while others, like qXR (Qure.ai, India) and Lunit INSIGHT CXR (Lunit Inc. Republic of Korea) also aim to replicate radiologist reports and claim to detect other lung findings in addition to TB (7).

Very few manufacturer-independent studies have assessed the diagnostic accuracy of CAD software for pulmonary TB (8–10); the limited number of studies, combined with methodological limitations, has hampered the development of guidance on the use of CAD thus far (10,11).

To enable the independent validation of CAD software products for their ability to detect pulmonary TB, FIND and partners have created a large library of digital CXR images, including clinical metadata, from individuals who were screened for TB. In parallel, the International Organization for Migration (IOM), established a global library of CXRs from migrants undergoing pre-migration TB screening in their resident country before migrating to the US.

The objective of both the FIND and IOM studies was to assess what the diagnostic accuracy is, in terms of sensitivity and specificity, of three commercially available, Confirmité Europëenne (CE)-certified CAD systems for detecting microbiologically confirmed TB in adults who are not seeking care but who are screened for TB in a health provider-initiated screening activity.

The analytic approach and CAD software versions were the same for both studies. In this paper, we describe the results using FIND’s global CXR TB screening library; the results of the IOM study using their migrant CXR TB screening library are described in a separate paper. The results of both studies were used in a guideline development group meeting convened by WHO in 2020 to inform an update of the TB screening guidelines, which resulted in the recommendation that CAD may be used as an alternative to human CXR interpretation for the purpose of TB screening (12,13).

## Methods

### Study population

FIND’s global CXR TB screening library contains digital CXR images (in DICOM, Digital Imaging and Communications in Medicine format) of individuals from different geographical regions and different risk groups that were taken as part of research studies or programmatic screening activities (henceforth called ‘source studies’). Source studies were considered eligible when all the following criteria were met: (1) CXR evaluation was carried out in a defined period for a cohort of individuals irrespective of the presence of symptoms; (2) CXRs were available in DICOM format and had not been previously been shared with any CAD developer for purposes of training their software; and (3) a minimal set of clinical (age, sex, co-morbidities) and laboratory variables was available, including microbiological test results from sputum samples. All reporting followed the STARD guideline (14) and a prespecified analysis plan was followed but was not officially published.

### Sample size

We aimed to include in our library a minimum of 590 CXRs for participants with TB (TB cases) and 868 for participants without TB (non-TB cases) according to the definitions of each of the reference standards. This sample size assumed a CAD system to be at least 90% sensitive and 70% specific, in line with the WHO target product profile (TPP) for a triage test in detecting pulmonary TB (16), and allowed a precision of the estimates expressed as total width of the 95% confidence interval (CI) of 10%, with 90% power of observing a CI of such width or less. The resulting sample size was increased by a factor of 10% to account for potential missing and/or incomplete data (17). The study was not powered to evaluate a difference between the performance of CADs and humans when using the same reference standard. The final sample size, however, allows to detect a difference of 10% in specificity (from a reference value of 45%) with power greater than 90%, as well as non-inferiority (for specificity of CADs when compared with human readers) with a margin of 10%.

### Sampling

From each source study, two sets of random samples were drawn based on different case definitions (Figure 1). Firstly, participants with a microbiological confirmation and classified according to the case definitions of the microbiological reference standard (MRS), described in **Table 1**, were sampled. For all studies except for the Philippines prevalence survey, the total number of microbiologically confirmed TB cases were included, given the relatively low number of TB cases. The Philippines prevalence survey contributed a sample of confirmed TB cases until the required number of confirmed TB cases was reached. For the sampling of non-TB cases, data providers were asked to randomly sample 1.5 times as many non-TB cases as TB cases from their study dataset to make sure that each study contributed a similar proportion of TB and non-TB cases to the final FIND library.

**Table 1.**
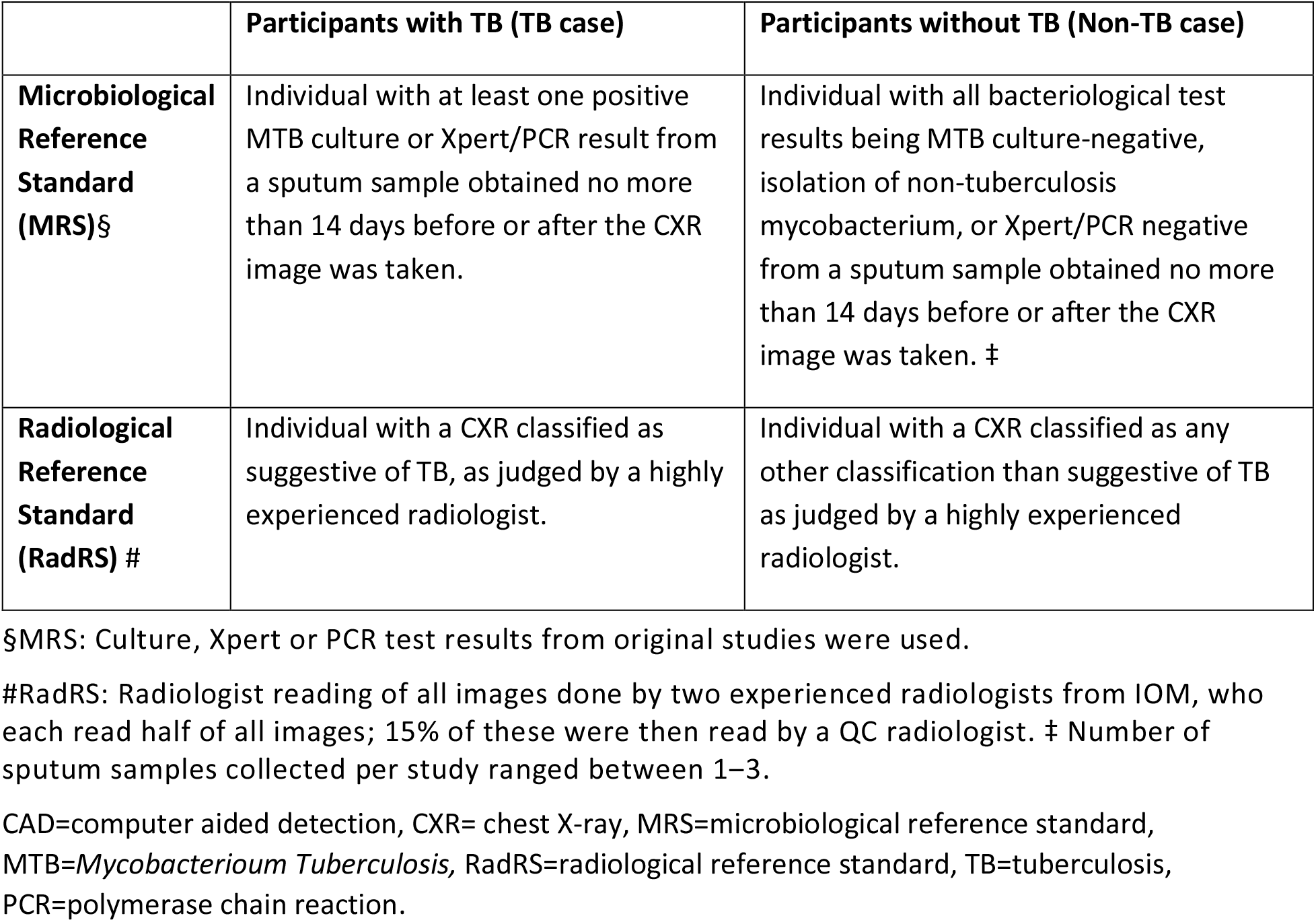
Definition of reference standards used in the evaluation of three CAD software products for chest X-ray interpretation in TB screening.

|  | Participants with TB (TB case) | Participants without TB (Non-TB case) |
| --- | --- | --- |
| **Microbiological Reference Standard (MRS)§** | Individual with at least one positive MTB culture or Xpert/PCR result from a sputum sample obtained no more than 14 days before or after the CXR image was taken. | Individual with all bacteriological test results being MTB culture-negative, isolation of non-tuberculosis mycobacterium, or Xpert/PCR negative from a sputum sample obtained no more than 14 days before or after the CXR image was taken. ‡ |
| **Radiological Reference Standard (RadRS) #** | Individual with a CXR classified as suggestive of TB, as judged by a highly experienced radiologist. | Individual with a CXR classified as any other classification than suggestive of TB as judged by a highly experienced radiologist. |

§MRS: Culture, Xpert or PCR test results from original studies were used.
#RadRS: Radiologist reading of all images done by two experienced radiologists from IOM, who each read half of all images; 15% of these were then read by a QC radiologist. ‡ Number of sputum samples collected per study ranged between 1–3.

**Figure 1.**
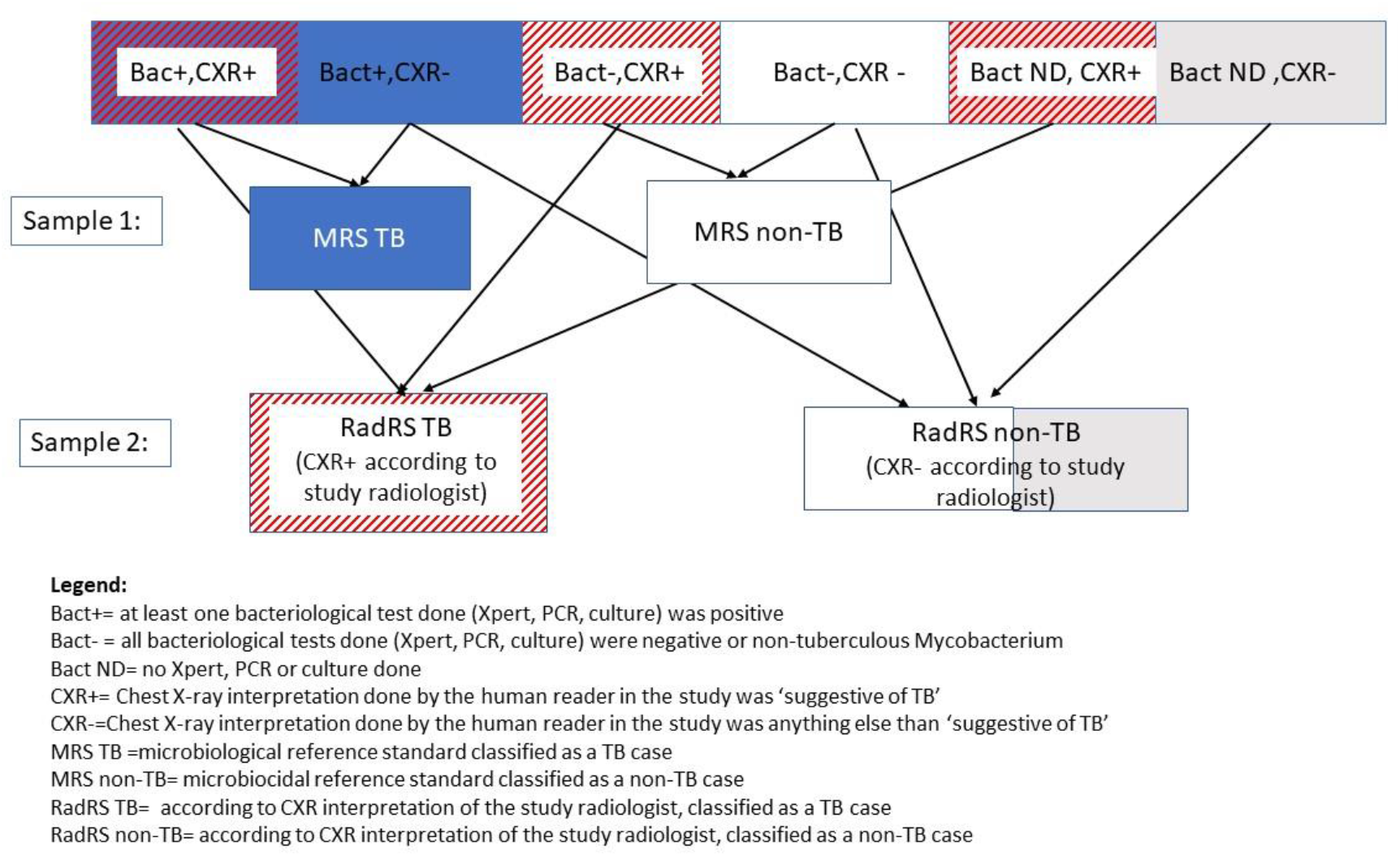
Flow diagram of the sampling procedure used in an evaluation of three CAD software products for chest X-ray interpretation in TB screening. CAD=computer-aided detection, CXR=chest X-ray, RadRS=radiological reference standard, MRS=microbiological reference standard, PCR=polymerase chain reaction, TB=tuberculosis.

The second sample that was drawn from each study drew the same number of TB and non-TB cases, but this time the case definitions were based on the interpretation of the CXR by the study radiologist. The two samples were combined. Overlap between the two samples was accepted, but any duplicates that were part of both samples were removed from the final library before analysis.

### Data preparation

The first DICOM image was selected for each study participant. If applicable, any identifying information was removed from the DICOM image (e.g., by use of a DICOM anonymizing tool or by overlaying a black square on the part of the CXR image containing personal data). Anonymized DICOM images and a dataset with subject demographics and clinical information were uploaded by the study teams to a secure server hosted at FIND headquarters in Geneva, Switzerland. All study subject datasets were then harmonized into one overarching dataset.

### Index test

We evaluated three commercially available CAD applications, all CE-certified before January 2020; CAD4TB v6 (Delft Imaging, Netherlands), qXR v2 (Qure.ai, India), and Lunit Insight CXR TB algorithm v4.9.0 (Lunit Inc., South Korea). All CAD applications were locally installed as offline software to ensure that the developers would have no access to the DICOMs in the library via their cloud. All CAD products gave a TB abnormality score, which ranged between 0 and 100 (for CAD4TB) or 0 and 1 (for qXR and Lunit). CAD analysis was carried out by FIND, and manufacturers only provided support through remote team viewer sessions when needed for trouble shooting. The manufacturer of CAD4TB had recommended to do a pre-validation for each X-ray system that was used, and provide a set of approximately 200 images (100 TB cases and 100 non-TB cases) to confirm that the software could correctly interpret the CXR images. Such pre-validation was not carried out because the amount of data needed from each site and from each X-ray system exceeded the number of available images from each study. Therefore, the restriction to only process DICOM images from pre-validated X-ray systems was lifted and CAD4TB was used off label for the FIND library.

### Modifications to DICOM images

The CAD4TB v6 software requires that DICOM images be taken as posterior-anterior (PA) CXR and saved vertically, and in monochrome 12-bit format. Since some images did not meet these specifications, modified DICOM versions were created when needed (see the Supplement for further details). Although qXR and Lunit did not require any such adjustments, all three CAD products were run on the same set of images, including the modified images, to allow an impartial comparison across the three products.

### Reference standards

The accuracy of the CAD products was evaluated against two reference standards (**Table 1**). For the microbiological reference standard (MRS), a TB case was defined as having at least one positive culture for *Mycobacterium tuberculosis* (MTB) or Xpert MTB/RIF (Cepheid, Sunnyvale, USA) or in-house positive polymerase chain reaction (PCR) result (henceforth summarized as ‘Xpert/PCR’) from a sputum sample obtained no more than 14 days before or after the CXR image was taken. A non-TB case was defined as all microbiological testing done (culture, Xpert/PCR) being negative or isolation of non-tuberculosis mycobacteria result from a sputum sample obtained no more than 14 days before or after the CXR image was taken. There was no restriction on the number of sputum samples that needed to be obtained per individual, so one sputum sample collected per study participant was considered sufficient. For the radiological reference standard (RadRS), a case suggestive of TB was defined as a CXR classified by an expert radiologist as either a) abnormal, highly suggestive of active TB (follow-up required), or b) abnormal, suggesting old, healed TB but active TB cannot be ruled out (follow-up required). A non-TB case was defined as a CXR suggesting any classification other than TB, which included images that were judged as a) normal; b) abnormal, remotely suggesting old, healed TB but minimal risk of reactivation (no follow-up required); c) abnormal, not suggestive of TB (follow-up required); or d) other abnormalities (no follow-up required). All CXRs were read by two certified expert IOM radiologists, each with over 6 years’ experience in the IOM health assessment program, and highly qualified in internal quality control. Each radiologist read a random set of half of the images using a standardized, prespecified CXR classification form. The radiologists were blinded to the initial CXR readings from the source study radiologists and the CAD scores, as well as the demographic and clinical information of the participants.

### Analysis

The distribution of the TB abnormality scores of the different CAD software in individuals classified as TB and non-TB was visually explored with histograms. Pooled receiver operating characteristic (ROC) curves were plotted for each CAD against each reference standard, pooling the data from the different source studies, and used to estimate the AUC of each CAD. The heterogeneity between source studies was explored at the study-level. Individual participant data meta-analyses as well as summary ROC (SROC) curves across the different source studies were subsequently computed to generate pooled estimates of the test accuracies while accounting for between-study heterogeneity, by use of a random effects bivariate model implemented with the use of the “mada” R package. Furthermore, the accuracy of tests in terms of sensitivity and specificity were evaluated at a set sensitivity of 90% and a set specificity of 70% (in line with WHO’s targets for a triage TPP), and at company-recommended thresholds for qXR and Lunit. Furthermore, the performance of the RadRS against MRS was determined in each screening source study dataset and a pooled, meta-analysed estimate was determined through use of a Bayesian hierarchical model (implemented in the R meta4diag package)(18). The performance of the CAD products was then compared at the same sensitivity and specificity as for the radiologists against MRS. Lastly, the AUC and sensitivity and specificity analyses were explored for pre-specified subgroups of participants. All analyses were performed using R v 4.03. Specific packages used for parts of this analysis include: pROC, mada, diagmeta and meta4diag (19).

### Ethics

All source studies contributing to the FIND CXR library obtained local ethical approval to allow sharing of the data. In addition, the study protocol for the ECAD-TB project was approved by the McGill University Health Centre International Review Board (protocol number 2019-4649).

## Results

This study was conducted between November 2018 and November 2021. Six screening source studies each contributed between 189 and 908 images for a total of 2,756 participants. A description of the six source studies and the demographic characteristics of the different study populations can be found in **Supplement Tables S1-S7**. Three source studies (studies 1, 2, and 3) had conducted targeted screening in high-risk groups: people living in congregate settings in a high prevalent area (XACT II in South Africa), people residing in correctional services (South Africa), or entry screening of asylum seekers (The Netherlands). The other source studies (studies 4, 5 and 6) had screened the general population as part of a country-wide prevalence survey (Vietnam (20), and Philippines (21)) or house-to-house screening in a province (22).

Out of the total 2,756 participants, 1,449 (53%) were classified as RadRS-positive and 1,307 (48%) as RadRS-negative. MRS results were available for 1,753 (64%) of the participants. All source studies, except the ACT-3X study, collected sputum samples only in participants who were symptomatic or had a CXR suggestive of TB and, therefore, MRS results were not available for 805 (29%) participants. 198 (7%) participants were excluded because the MRS test results and CXRs were done more than 14 days apart. Of the 1,753 participants with MRS results, 528 (30%) were MRS-positive and classified as TB, and 1,225 (70%) MRS-negative and classified as non-TB (**Figure 2**). For the remainder of this paper, we concentrate on the results against MRS. The same analysis as presented for MRS was performed against RadRS (see **Supplement Tables S9-S12, Figures S4-S6**).

**Figure 2.**
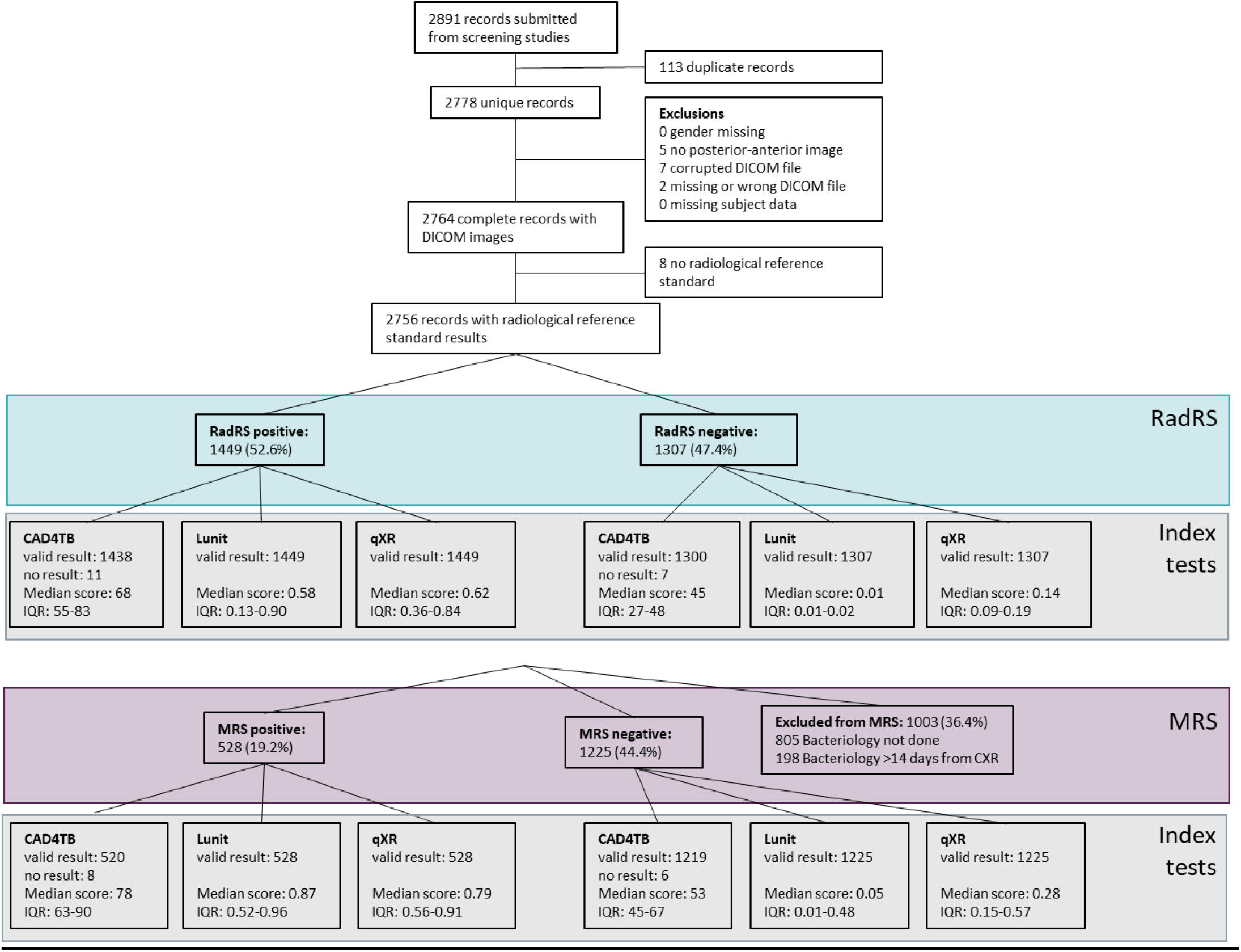
Flow diagram of an evaluation of three CAD software products for chest X-ray interpretation in TB screening, showing the number of eligible participants with a screening DICOM image included in the accuracy analysis against the microbiological reference standard. Legend: CAD=computer-aided detection, CXR=chest X-ray, DICOM= Digital Imaging and Communications in Medicine – digital CXR image, IQR=inter quartile range, MRS=microbiological reference standard, RadRS=radiological reference standard

Most participants with MRS results originated from the WHO-classified Western Pacific Region (68%) and the African region (28%) (**Table 2**). Over half of the participants were male (65%) and the median age was 47 (IQR 34-61) years. TB symptoms were present in 968 (55%) of the participants in our library, with an almost equal proportion of symptomatic participants among those classified as MRS-positive (59%) and MRS-negative (54%). A known history of TB was present in 295 (17%) of all participants and 137 (8%) were known to be HIV-positive. CXRs were judged as abnormal by the expert radiologist in 494 (94%) of the MRS-positive cases but also in 611 (50%) of the MRS-negative ones.

**Table 2.**
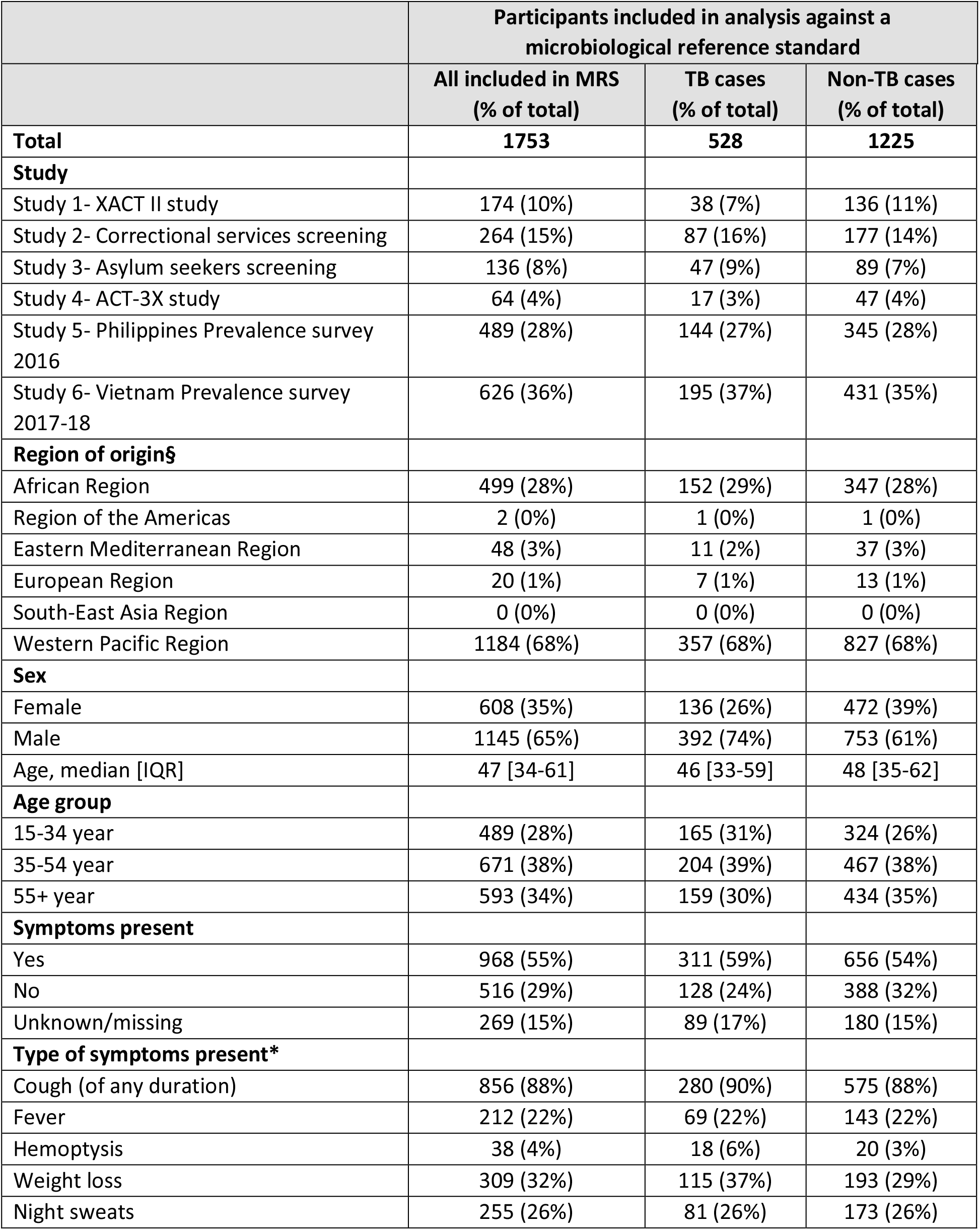
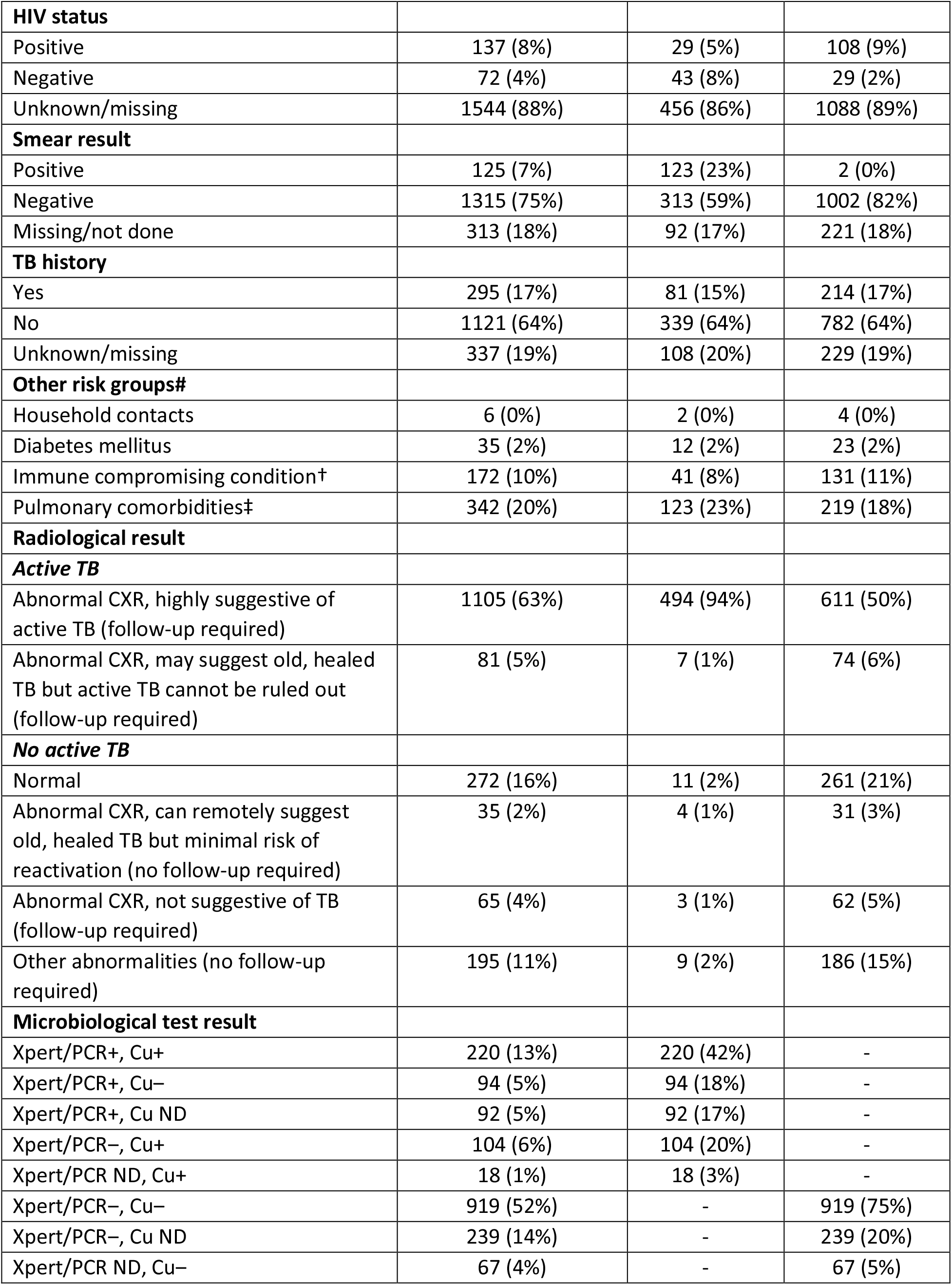
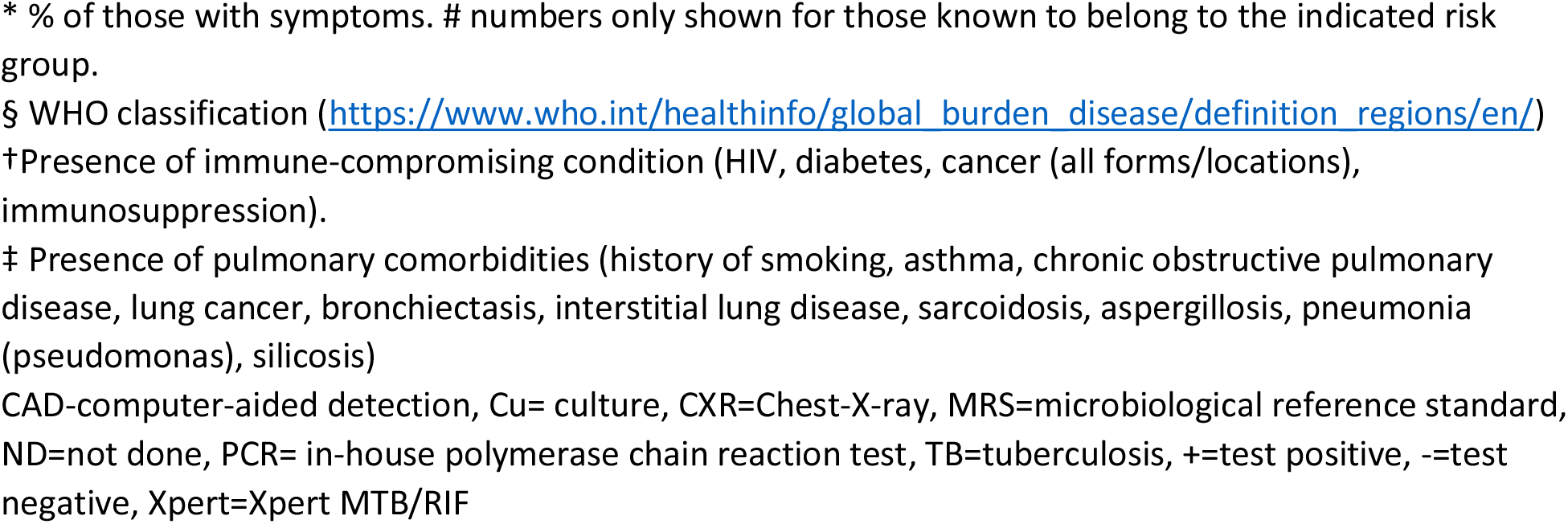
**Demographic and clinical characteristics of participants included in an evaluation of the diagnostic accuracy of three CAD software products for chest X-ray interpretation against a microbiological reference standard in TB screening**

Five out of six source studies had conducted culture, Xpert/PCR and sputum smear testing, whereas one (study 2) had only done Xpert/PCR. Of all microbiologically-confirmed TB cases, 342 (65%) had a positive MTB culture result, 92 (17%) had a positive Xpert/PCR result but had no culture testing done, and 94 (18%) had a positive Xpert/PCR result but negative culture result.

### Distribution of abnormality scores

All DICOM images had an abnormality score generated by qXR and Lunit; for 18 images (14 with MRS results) there was no valid CAD4TB result, with images excluded from the statistical analysis for CAD4TB only. For all three CAD products, median scores were higher in TB than in non-TB classified images, irrespective of which reference standard was used, with the highest median scores found in the MRS TB group (**Figure 2**). All CAD abnormality scores followed a bimodal distribution, generally with lower CAD scores in non-TB cases and higher abnormality scores among TB cases, although the CAD score distributions in TB cases and non-TB cases were overlapping. For CAD4TB, the two modes for non-TB and TB cases were closer to each other, while for Lunit and qXR the two modes were closer to the opposite extremes (**Supplement Figure S1**).

### Accuracy estimates

ROC curves plotted against the MRS showed that the performance of CADs varied across study sources (**Supplement Figure S2**). While the area under the curve (AUC) was highest for Lunit in 6 out of 7 studies, CIs were wide and overlapped with those of CAD4TB and qXR for all the studies (**Table 3**).

**Table 3.** **Area under the receiver operating characteristic (ROC) curve of the performance of three CAD software products for chest X-ray interpretation in each contributing TB screening study as well as pooled ROC measured against a microbiological reference standard**

|  | Number of participants in analysis |  | CAD4TB | Lunit | qXR |
| --- | --- | --- | --- | --- | --- |
|  | TB | Non-TB | AUC (95% CI) | AUC (95% CI) | AUC (95% CI) |
| Study 1- XACT II study | 38 | 136 | 67.0 (56.9-77.1) | 69.8 (59.0-80.6) | 66.4 (55.6-77.1) |
| Study 2- Correctional services screening | 87 | 177<sup>#</sup> | 74.5 (68.3-80.8) | 83.3 (77.9-88.7) | 79.3 (73.6-84.9) |
| Study 3- Asylum seekers screening | 47 | 89 | 65.7 (55.9-75.6) | 81.6 (74.2-89.0) | 76.8 (68.3-85.2) |
| Study 4- ACT-3X study | 17 | 47 | 85.4 (74.0-96.8) | 86.9 (74.9-98.8) | 88.7 (77.4-99.9) |
| Study 5- Philippines Prevalence survey 2016 | 144<sup>Ω</sup> | 345<sup>§</sup> | 83.5 (79.5-87.6) | 90.0 (87.0-93.0) | 86.8 (83.2-90.3) |
| Study 6- Vietnam Prevalence survey 2017-18 | 195<sup>^</sup> | 431<sup>~</sup> | 78.3 (74.6-81.9) | 84.0 (80.8-87.2) | 80.7 (77.2-84.2) |
| **Simple pooled ROC** | **528<sup>†</sup>** | **1225<sup>‡</sup>** | **77.8 (75.4-80.2)** | **84.1 (82.1-86.1)\*** | **81.2 (79.0-83.4)** |
| **Summary ROC** | **528<sup>†</sup>** | **1225<sup>‡</sup>** | **76.4 (72.1-80.3)** | **83.3 (78.4-87.2)** | **80.4 (75.9-84.2)** |

AUC=area under the curve, CAD=computer-aided detection, simple pooled ROC =simple pooled ROC curve combining all screening studies.
<sup>#</sup> Non-TB: n=176 for CAD4TB (1 participant had missing CAD4TB results)
<sup>Ω</sup> TB: n=137 for CAD4TB (7 participants had missing CAD4TB results)
<sup>§</sup> Non-TB: n=341 for CAD4TB (4 participants had missing CAD4TB results)
<sup>^</sup> TB: n=194 for CAD4TB (1 participant had missing CAD4TB results)
<sup>~</sup> n=430 for CAD4TB (1 participant had missing CAD4TB results)
<sup>†</sup> n=520 for CAD4TB (8 participants had missing CAD4TB results)
<sup>‡</sup> n=1219 for CAD4TB (6 participants had missing CAD4TB result)
\* 95% CI of AUC for Lunit is not overlapping with that for CAD4TB and qXR

When pooling the data from all studies, AUCs from the summary ROC curves of the meta-analysis were 83.3 (95% CI 78.4-87.2) for Lunit, 80.4 (95% CI 75.9-84.2) for qXR, and 76.4 (95% CI 72.1-80.3) for CAD4TB but with overlapping CIs for all three products (**Figure 3**).

**Figure 3.**
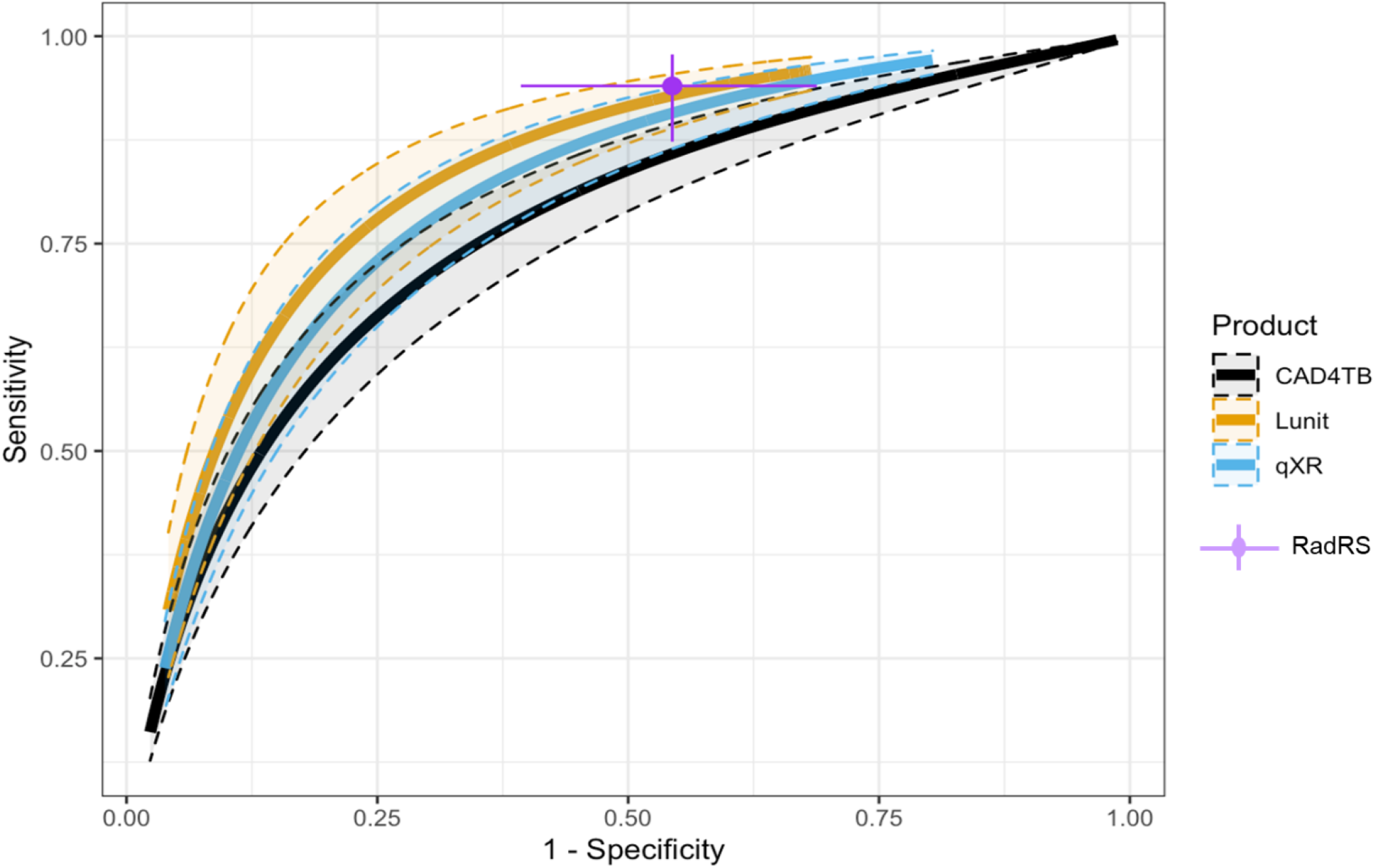
Summary ROC curves illustrating the performance of three different CAD software products for chest X-ray interpretation against a microbiological reference standard (MRS) in a TB screening population. Legend: The colored lines represent the estimated summary ROC curve as predicted by the R diagmeta function for CAD4TB (black line), Lunit (orange) and qXR (blue). The shaded area around each of the plotted lines represents the 95% confidence interval (CI) indicating the uncertainty around the estimated summary ROC curve. The purple dot shows the accuracy of the radiologist reference standard (RadRS) and 95% CI, when compared against MRS, which corresponds to a sensitivity of 94.0 (95% CI 87.3−97.8) and specificity of 45.6 (95% CI 31.2−60.7). CAD=computer-aided detection, ROC=receiver operating characteristic, TB=tuberculosis

We estimated the performance of the expert radiologists (RadRS) against MRS, which resulted in an estimated sensitivity of 94.0% (95% CI 87.3−97.8), with a specificity of 45.6% (95% CI 31.2−60.7). At the same sensitivity of 94.0%, all CAD software had lower point estimates for specificity than the radiologists (22.4%, 95% CI 16.9-29.0 for CAD4TB, 34.8%, 95% CI 25.3-45.1 for qXR and 41.0%, 95% CI 30.1-53.0 for Lunit, **Table 4**). For qXR and Lunit, the CIs overlapped with those of the radiologists, suggesting that the software could be performing similarly, while CAD4TB had a significantly lower specificity compared to the radiologist and Lunit. At a set specificity of 45.6%, all CADs had again lower point estimates for the sensitivity but overlapping CIs with the sensitivity estimate of the radiologist.

**Table 4.** **Predicted sensitivity and specificity estimates for three different CAD software products for chest X-ray interpretation against a microbiological reference standard at various threshold scores for each CAD based on the summary ROC curves in a TB screening population.**

|  | Software | Sensitivity (%)<br>(95% CI) | Specificity (%)<br>(95% CI) | Threshold<br>score# |
| --- | --- | --- | --- | --- |
| Set sensitivity of<br>90% | CAD4TB | 90.0 (86.5–92.7) | 34.9 (27.9–42.8)<sup>∞</sup> | 45.8 |
|  | Lunit | 90.0 (84.9–93.5) | 54.5 (43.0–65.6) | 0.24 |
|  | qXR | 90.0 (85.4–93.2) | 47.7 (37.5–58.6) | 0.31 |
| Set specificity of<br>70% | CAD4TB | 71.0 (64.4–76.9) | 70.0 (63.4–75.9) | 63.3 |
|  | Lunit | 82.0 (74.4–87.8) | 70.0 (59.9–78.5) | 0.41 |
|  | qXR | 77.6 (70.3–83.6) | 70.0 (61.6–77.3) | 0.49 |
| Developer<br>recommended<br>thresholds (thr) | CAD4TB | – | – | None |
|  | Lunit – thr1 | 92.9 (89.0–95.5) | 45.4 (34.0–57.2) | 0.15 |
|  | Lunit – thr2 | 87.8 (81.9–92.0) | 59.9 (48.6–70.3) | 0.30 |
|  | Lunit – thr3 | 79.8 (71.6–86.1) | 72.9 (63.3–80.7) | 0.45 |
|  | qXR – thr 1 | 72.2 (64.2–79.0) | 75.6 (68.3–81.7) | 0.55 |
|  | qXR – thr 2 | 48.4 (40.3–56.5) | 89.5 (86.0–92.2) | 0.75 |
| Same sensitivity as<br>radiologists (RadRS)<br>*[TB=active TB or old*<br>*healed TB but*<br>*active TB cannot be*<br>*ruled out]* | RadRS | 94.0 (87.3–97.8) | 45.6 (31.2–60.7) |  |
|  | CAD4TB | 94.0 (91.6–95.7) | 22.4 (16.9–29.0)<sup>†</sup> | 38.4 |
|  | Lunit | 94.0 (90.6–96.2) | 41.0 (30.1–53.0) | 0.11 |
|  | qXR | 94.0 (90.9–96.1) | 34.6 (25.3–45.1) | 0.20 |
| Same specificity as<br>radiologists (RadRS) | RadRS | 94.0 (87.3–97.8) | 45.6 (31.2–60.7) |  |
|  | CAD4TB | 85.9 (81.4–89.4) | 45.6 (37.9–53.6) | 51.1 |
|  | Lunit | 92.8 (88.9–95.4) | 45.6 (34.3–57.4) | 0.15 |
|  | qXR | 90.7 (86.4–93.8) | 45.6 (35.5–56.1) | 0.29 |

Sensitivity and specificity estimates were assessed at different thresholds for the abnormality score. At a set sensitivity of 90%, the specificity was 34.9% (95% CI 27.9-42.8) for CAD4TB, 47.8% (95% CI 37.5-58.6) for qXR and 54.5% (95% CI 43.0-65.6) for Lunit, with non-overlapping CIs between CAD4TB and Lunit. At a set specificity of 70%, sensitivity was 71.0% (95% CI 64.4-76.9) for CAD4TB, 77.6% (95% CI 70.3-83.6) for qXR and 82.0% (95% CI 74.4-87.8) for Lunit, with all having overlapping CIs. The thresholds recommended by the developers of Lunit all had relatively high sensitivity (with respective sensitivity and specificity of 92.9% and 45.4% at threshold 0.15, 87.8% and 59.9% respectively at threshold 0.30, and 79.8%, and 72.9% at threshold 0.55) while those of qXR had relatively higher specificity (with respective sensitivity and specificity of 72.2% and 75.6% at threshold 0.55 and 48.4% and 89.5% at threshold 0.75).

### Subgroup analysis

Subgroup analysis indicated that the performance of CAD may be reduced in female participants, in individuals with a history of TB, and in people without symptoms (**Figure S3**). However, CIs around AUC estimates were wide and overlapping for all subgroup comparisons against MRS.

### Comparison of CADs against the radiological reference standard

Compared against RadRS, a comprehensive analysis is presented in **Tables S9-12** as well as in **Figures S4-S6** in the **Supplement**. In brief, all 3 CAD programs had higher estimates of AUCs when compared to MRS across the different screening datasets, as well as in the pooled meta-analyzed SROC analysis: 87.8 for Lunit (95% CI 85.7−89.3), 88.0 for CAD4TB (95% CI 85.9-89.9) to 90.5 for qXR (95% CI 87.8-92.7) (**Table S10, Figure S5, Figure S6**).

Accuracy estimates at different threshold scores were higher in most comparisons for all products when evaluated against RadRS. At a set sensitivity of 90%, specificity was 64.5% (95% CI 58.9-69.7) for CAD4TB and 66.5% (95% CI 59.0-73.1) for qXR but 44.3% (95% CI 40.0-48.7) for Lunit, which was significantly lower than for the other two products (**Supplement Table S6**). At a specificity of 70%, sensitivity reached 87.9% (95% CI 85.2-90.2) for CAD4TB, 86.1% (95% CI 83.7-88.2) for Lunit, and 89.3% (95% CI 86.0-92.0) for qXR.

## Discussion

In this individual participant data meta-analysis of three different commercial CAD products used to test for TB in a screening population, which we believe is the first of its kind, we showed that the accuracy measured against a microbiological reference standard is reasonable and may achieve a sensitivity and specificity that approximates those of experienced radiologists in identifying individuals who require further confirmatory testing for TB. Moreover, all three CAD products had a high and largely comparable accuracy when compared against a radiological reference standard of experienced radiologists. While there was variability in the performance of the CADs across the different source studies and small differences existed between the software within each study, the meta-analyzed pooled AUCs of the three CADs were similar when assessed against MRS or RadRS.

However, none of the CAD products or the expert radiologists in our study met the aspirational targets set by WHO for a triage test. There were also some differences between the products. When the specificity of the CADs was compared at a set sensitivity of 90%, CAD4TB had the lowest specificity at 34.9% compared to 47.7% for qXR and 54.5% for Lunit using MRS, while Lunit had a lower specificity (44.3%) compared to CAD4TB (64.5%) and qXR (66.5%) when using a RadRS. While qXR and Lunit had a slightly lower yet not statistically different specificity than that of the expert radiologist’s, the specificity of CAD4TB was significantly lower than that of the expert radiologist when their sensitivity was matched,.

Our screening archive contained data from a variety of risk groups that are recommended for targeted TB screening given their high TB prevalence. Very few peer-reviewed studies have assessed the performance of CADs in such groups, and all studies except one (23) used earlier versions of CAD4TB (24–26). Two of these studies suggested that CAD4TB v5 or v6 could achieve a similar performance as that of a trained field radiologist when interpreting CXRs of the Zambia National TB prevalence survey (25), or a senior radiologist when interpreting images from a population-based survey in a high HIV-endemic area in rural South Africa (23). But the accuracy of the field radiologists in both of these studies had a sensitivity of 69.8% (with a specificity of 92.6%) and 80.8% (with a specificity of 66.9%), values considerably lower than the sensitivity of the expert radiologists in our study, which was 94% (with a specificity of 45.6%) (23,25).

CXR examination with CAD interpretation has the potential to make screening programs more efficient, especially when done at scale, given their ability to generate instantaneous, standardized results on an endless number of CXRs without the reader fatigue or inter-reader variability that human readers face. It is to be expected that, in settings where X-ray devices are available but access to human readers is limited or lacking, the use of CADs may substantially increase the population that can be reached for testing and increase the number of individuals who receive confirmatory diagnosis and initiate treatment; implementation and cost-effectiveness studies are needed to demonstrate how and when the rollout of CAD will have the most impact in real-life settings. Recent advances in X-ray technology and the development of ultra-portable digital X-ray devices may make TB CXR screening in remote or hard to reach areas and for sub-populations a realistic target (27). Nevertheless, cost of equipment and artificial intelligence software licences are high and still a major barrier for large scale-up without substantial donor support (27,28).

The performance of CAD in terms of sensitivity and specificity at a given threshold score varied across studies, meaning that the same abnormality threshold score of a particular CAD product has a different sensitivity and specificity in each setting. Because of this, and before implementation of a CAD product, WHO recommends implementers conduct a calibration study to determine the most appropriate threshold score for their setting and use-case. The Special Programme for Research and Training in Tropical Diseases has developed an online toolkit that allows implementers to model cost-effectiveness and impact based on the data collected in a calibration study (29).

### Strengths and limitations

Our study has several strengths. It is the largest combined validation dataset of screening CXR images from different geographical regions and different screening groups. None of the images have been used for the training of CAD products. At the same time, none of the data were specifically collected for the purpose of validating CAD products. Nevertheless, to date, this is the most representative (global) dataset for CAD validation that was built with the intention to use in future independent validations of CAD solutions.

It is important to note that we did not limit our analysis to a pre-specified threshold score as was recommended in the opinion paper by Khan *et al* (10). Before implementation, the CAD4TB manufacturer’s advice was to conduct a verification process to confirm that the software performance (AUC) was acceptable and could set or endorse a threshold. We did not carry out such a verification process for any of the CAD software because the number of images required (from each data provider) exceeded the number of images available for these studies.

While our dataset addresses many of the methodological shortcomings that were raised during the 2016 WHO meeting (10,11), including issues around study population, participant selection, and reference standards, there are still several possible reasons why the performance of the radiologists versus the CAD software is potentially underestimated or overestimated: (1) MRS analysis for the screening use-case was limited to individuals from whom a sputum sample was collected and in whom the presence of symptoms or an abnormal CXR were often the main reason for referral, thereby excluding healthy, asymptomatic individuals who are likely not to have TB. The exclusion of this group may potentially have resulted in an overestimation of the sensitivity rate (since most TB cases had sputum done because of an abnormal CXR) and underestimation of the specificity rate. The impact on the performance would be similar for the radiologist as well and thus the comparison of CADs against the RadRS would be less biased. (2) The inclusion of study 1 could have negatively affected the sensitivity estimate in the screening use-case, as HIV-negative participants in this study were only referred for CXR when they had a positive microbiological test result. Moreover, the inclusion of study 2, in which Xpert testing was only done after an initial CAD4TB v5 score of 60, may also have introduced bias. (3) Another limitation was that our analysis was based on retrospective data from different studies and processes were not controlled or standardized across studies. This might have resulted, for example, in culture results being variable between sites, with over decontamination resulting in false-negative results and thus lower specificity for the radiologist and CAD solutions when compared against the MRS. (4) Lastly, our analysis did not have enough data to confirm whether the accuracy of CAD solutions was impacted by certain risk factors, as has been suggested by Tavaziva *et al* (30). In their recent individual patient data meta-analysis of triage studies, the authors found that for the same CAD products, the sensitivity was modified by smear status and HIV infection and the specificity is significantly modified by age and prior TB.

In conclusion, we showed that three commercially available CAD products perform reasonably well in identifying individuals with CXR findings suggestive of TB and who require further confirmatory testing as part of TB screening, with a sensitivity and specificity that approximates those of experienced radiologists. Based on the results from our study, the companion study at IOM, and data presented by others (Qin *et al*, (personal communication), (30)), WHO is now recommending that in individuals aged 15 years and older, CAD may be used in place of human readers for interpreting digital CXR for screening for TB disease.

## Data Availability

Data cannot be shared publicly because FIND would like to re-use the DICOM library for future independent performance assessments of new AI versions or products to inform WHO policy updates and therefore the content of the data cannot be made available to AI developers in this field.

## Acknowledgements

We express our thanks to all participants who took part in the studies and screening projects, as well as to all teams at the participating study sites that contributed data from their completed studies or projects for their invaluable time and efforts in data preparation.

We also thank the developers Delft Imaging, Qure.ai and Lunit Insight for their support in providing their CAD products, carrying out the installation, and for technical support in troubleshooting issues and answering questions.

We are grateful to the IOM Global Teleradiology and QC Center technical team, Mr. Bhaskar Amatya, for their guidance, and IOM radiologists Drs. Mark Anthony Sy and Karla Francesca Zialcita who did the re-reading of the chest X-rays, and with Drs. Tesfa Semagne Egzertegegne and Lena Maria Ablis-Sun for their quality control reviews of the chest X-rays.

Finally, we thank Drs. Cecily Miller and Dennis Falzon from the WHO TB programme for their technical collaboration during the study and allowing us to present results of the studies at the WHO TB guideline development group meeting.

## Disclaimer

The findings and conclusions of this report are those of the authors and do not necessarily represent the official position of the US Centers for Disease Control and Prevention. References in this manuscript to any specific commercial products, process, service, manufacturer, or company does not constitute its endorsement or recommendation by the U.S. Government or CDC.

## Related manuscripts

This study done at FIND is one of the two studies on accuracy of CAD in a screening use case which were conducted at FIND and IOM under the same umbrella study protocol but using different data source and specific methods. The study conducted at IOM is being submitted separately for publication.

## Authors’ contributions

SVK contributed to the conceptualization, data curation, analysis, design of methodology, interpretation of the data, and writing of the initial draft of the manuscript.

SMG contributed to the conceptualization, design of methodology, interpretation of the data, and review and editing of the manuscript.

MR, OG, NM contributed to the interpretation of the data and review and editing of the manuscript.

RS, FAK, KL, FC, AB, PD contributed to the conceptualization, design of methodology, interpretation of the data, and review and editing of the manuscript.

RvH, VC, NVN, AE, AMCG, GBM, OA, KM, LTNA, KD, GJF contributed to the data curation, interpretation of the data, and review and editing of the manuscript.

SO, CMD contributed to the conceptualization, analysis, design of methodology, interpretation of the data, and review and editing of the manuscript.

All co-authors reviewed and agreed on the final manuscript.

